# Requirements to minimize airborne infections related to virus aerosol contamination at indoor cultural events

**DOI:** 10.1101/2022.11.07.22281932

**Authors:** Tunga Salthammer, Heinz-Jörn Moriske

## Abstract

The SARS-CoV-2 pandemic has resulted in many live events being canceled or held without spectator participation. It is therefore necessary to develop strategies to determine the conditions under which cultural activities can be maintained. In this study the results from available literature were combined with findings, guidelines and regulations for other indoor environments and recommendations were derived. In the cultural sector, the number of experimental investigations, surveys and simulations is comparatively small. This is probably due to the complexity of the events in terms of location and visitor flow, so the respective conditions under which they take place can be very different. It is therefore practically impossible to predict the risk of infection for a specific situation with potential virus spreaders attending or to derive general rules that go beyond the known measures of vaccination, testing, masks and distance. Cultural events can be held under pandemic conditions, provided certain conditions are met. Most study results agree on this. However, any recommendations for hygiene, safety and ventilation measures in cultural institutions can only minimize the risk of infection, but cannot completely rule it out. It is also of considerable importance that visitors protect themselves individually and act responsibly.

**Graphical Abstract:** 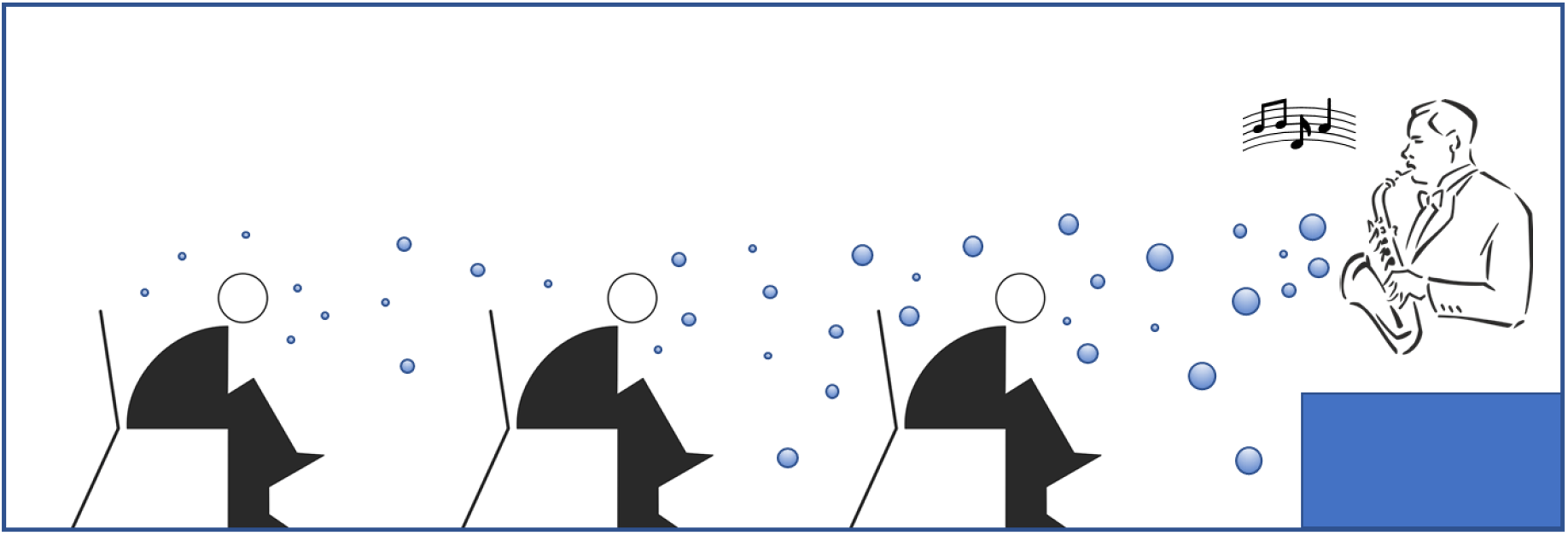

## Introduction

The cultural sectors in Europe are among the most diverse in the world. Art and culture are an integral part of society. Above all, cultural events contribute to social cohesion in their function as places of encounter and participation. The corona pandemic and the associated standstill in large parts of the art and culture scene has made it particularly clear how much poorer our society is without the direct experience of art and culture. In view of the current situation and with a view to possible future waves of infection and pandemics, efforts must be made to make cultural events as pandemic-proof as possible. The operation of opera houses, theatres, concert halls, cinemas and other venues should be maintained as long and as far as possible without having to compromise on infection protection.

For this purpose, the establishment of a uniform, professionally recognized and certified hygiene standard for cultural institutions makes sense. This should contain comprehensible and transparent criteria for infection protection that are relevant for the operator as well as for the staff and visitors. In addition, such a standard can serve as a basis for decision-making for the federal states and their authorities responsible for infection protection.

The present overview study creates a basis for the intended hygiene standards and ensures that these correspond to the current state of science and research. In particular, it summarizes the current state of knowledge with regard to the technical requirements for drastically minimizing aerosol-based infections at indoor cultural events and gives recommendations for various measures with a focus on ventilation, air distribution, air purification and CO_2_ monitoring in cultural institutions with regard to the risk of infection by SARS-CoV-2. However, the influence of the vaccination status will not be considered. General Information on the physical properties of bioaerosols, the risk of infection, and the use of face masks is briefly summarized, as this is necessary for understanding the available literature and the proposed measures.

As of March 2022, the recommendation for uniform hygiene and ventilation measures for cultural institutions under pandemic conditions and during normal operation, which was drawn up by an expert committee on the initiative of the Federal Government, is already available [1], which can be accessed as of August 10, 2022 at https://www.bundesregierung.de/breg-de/themen/buerokratieabbau/fuer-einheitlichere-corona-regeln-in-der-kultur-2010308.

## Methods

The usual databases (SCOPUS, Web of Science, PubMed and GoogleScholar) were consulted. The keywords or keyword combinations “SARS-CoV-2”, “Covid-19” and “Indoor” resulted in around 3400 hits. With a refined keyword search (“cultural”, “event”, “concert”, “singing”, “choir”, “ventilation”, “face mask”), the number of potentially interesting publications was limited to 367, although only a small proportion directly related to the risk of infection at cultural events. No studies were taken into account that deal with the economic situation of the cultural scene and venues or with the mental state of artists. As far as possible, the findings from other studies, for example in school classrooms [2], were applied to the recommendations for this topic. The flow chart according to PRISMA [3] of the systematic literature selection is shown in Figure 1. Recommendations for the cultural sector were then derived on the basis of the available literature.

**Figure 1.**
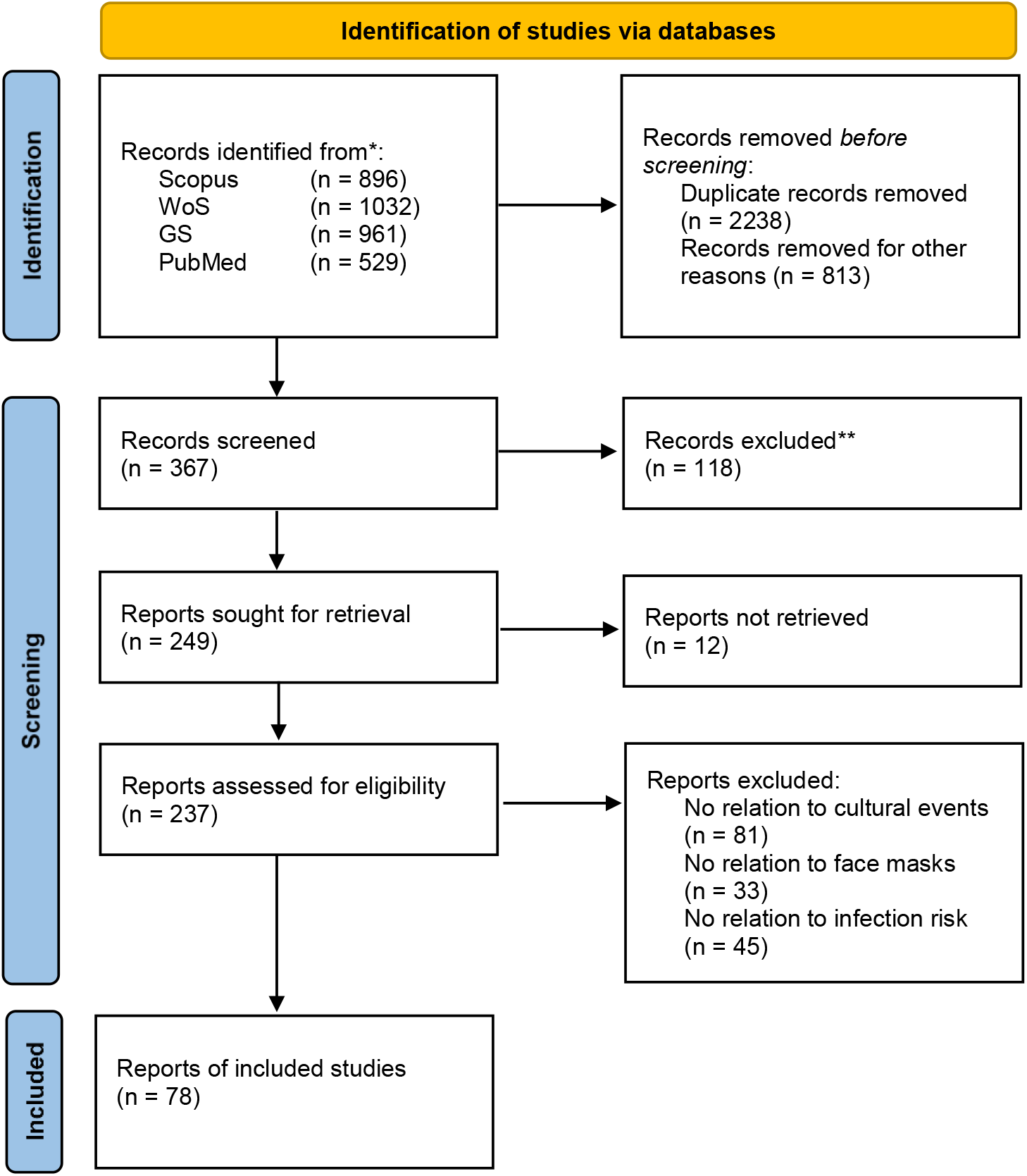
PRISMA flow diagram according to Page et al. [3] for the systematic review which included searches of databases. See also https://prisma-statement.org/PRISMAStatement/FlowDiagram, WoS = Web of Science, GS = GoogleScholar.

### Properties of Bioaerosols

Today it is undisputed that the respiratory intake of bioaerosols that arise when breathing, coughing, speaking and sneezing is one of the main transmission routes for SARS-CoV-2 viruses [4, 5]. From a physical point of view, an aerosol is a heterogeneous mixture of particles together with the gas or gas mixture surrounding them. In a stable aerosol, the liquid or solid components are homogeneously distributed as floating particles. Everyone exhales liquid particles of different sizes. If a person is infected with a pathogen, these particles can also contain viruses or bacteria, which become airborne and are inhaled by other people.

Bioaerosols are accumulations of particles in the air, which contain fungi (spores, conidia, fragments of hyphae), bacteria, viruses, pollen and their cell wall components and metabolic products (e.g. endotoxins, mycotoxins). Bioaerosols usually have aerodynamic diameters in the size range between 0.01 µm and 100 µm. SARS-CoV-2 is a membrane-enveloped RNA virus with a size between 60 nm and 140 nm. In the air, however, it is usually surrounded by a watery envelope. Under laboratory conditions, viable viruses have been found to be detectable in airborne aerosols up to 3 hours after release [6].

Particles with a diameter of approximately 10 µm deposit within minutes and particles with a diameter of approximately 100 µm within seconds in still air. However, in real environments, the particles are also transported by air movement (advection and turbulent transport) and can therefore remain in the air for much longer. Aqueous particles, however, evaporate in the air depending on their size, temperature and humidity. Figure 2 shows the time it takes for aqueous particles of different sizes to halve their diameter through evaporation at 20 °C and different relative humidities. For comparison, the time required for a particle to sink by 1 m in still air (black curve) is also plotted. The calculation was carried out according to Hinds [7]. The diameter of a 100 µm particle is therefore halved by evaporation in dry air within 5 seconds and in humid air (90% relative humidity) in about 1 minute. The evaporation time of a drop of water in still air is a function of the particle diameter, the particle density, the temperature of the particle and the ambient air, and the vapor pressure. This means that even large droplets do not necessarily fall to the ground but evaporate beforehand and remain in the air. At 20 °C and 50% RH, the evaporation time of 80 µm particles is of the order of the falling time (see Figure 2).

**Figure 2.**
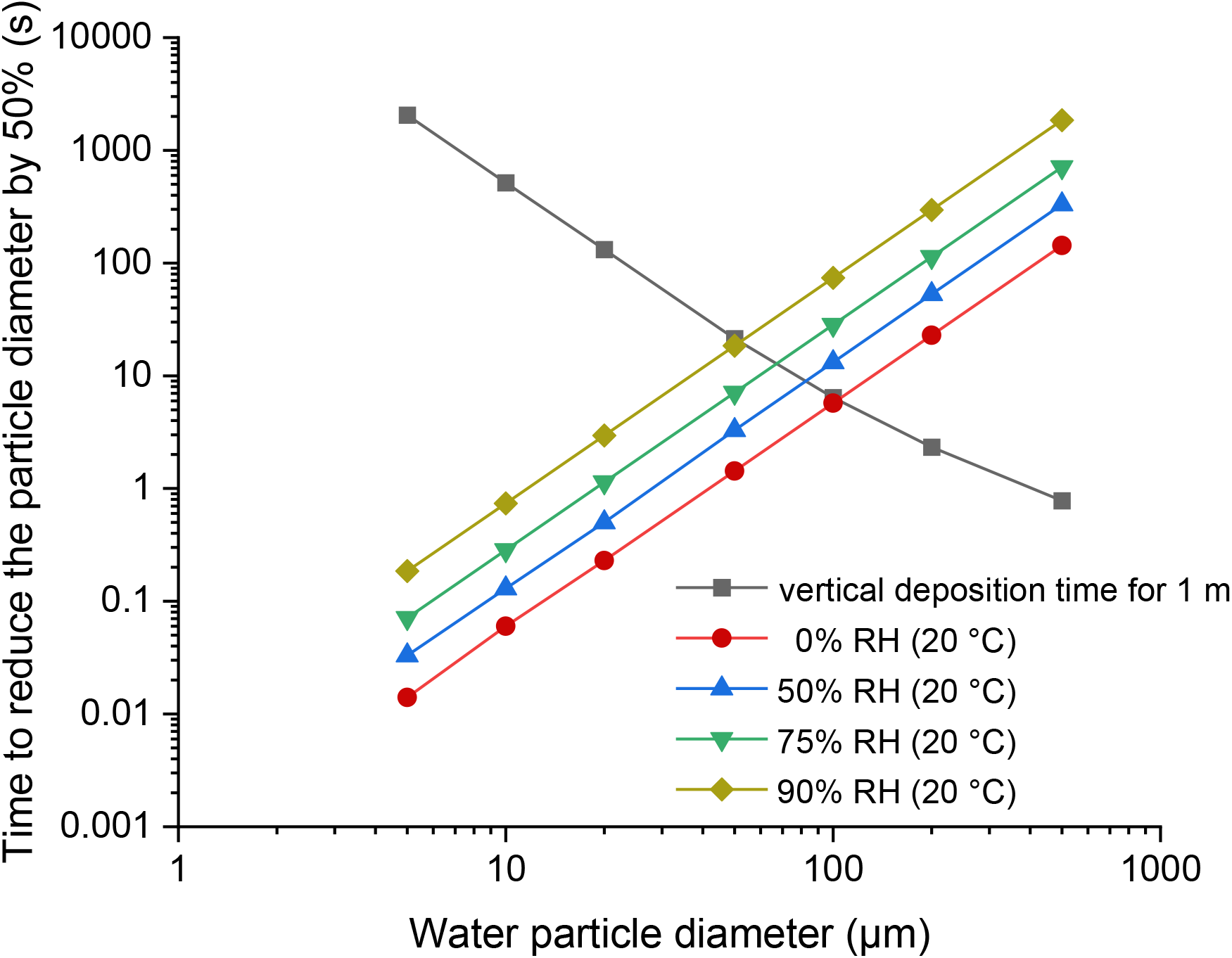
Time required for the diameter of aqueous particles to decrease by 50% through evaporation at 20 °C and various relative humidities. For comparison, the time required for a particle to sink by 1 m in still air is also plotted (black curve).

A very good overview of SARS-CoV-2 related aerosol physics can be found in the position paper of the Gesellschaft für Aerosolforschung (GAeF), authored by Asbach et al. [8].

### Properties of exhaled particles

The composition of the aerosol exhaled by humans depends on the respective activity. Speaking softly exhales fewer particles and droplets than speaking, singing, and shouting loudly. In addition, the ejection velocities are very different. The highest velocities are reached when coughing and sneezing. The number and size of aerosol particles and droplets released during human activities such as breathing, speaking, coughing and sneezing have been well studied, and the results have been published in various studies.

In one study, the release of droplets was measured while counting from 1 to 100. It was found that the number of particles had a maximum at 1 µm [9] and 6 µm [10]. In another study, the size range from 45 µm to 150 µm was examined [11]. Depending on the volume, emission rates of 1 to 50 particles were measured, but the particle size distribution was independent of the volume. Some individuals are “super-emissive” and emit about an order of magnitude more particles than average [9]. Edwards et al. [12] found that normal emitters have 14 - 71 particles/l in exhaled air, while superemitters have around 660 - 3230 particles/l.

Droplets produced by coughing have been found to exhibit two particle modes. Loudon and Roberts [13] report that 466 particles are released per cough reflex. The sizes of the droplets were distributed between 2 μm and 1471 μm, with maxima at 2 μm and 26 μm [14]. Xie et al. [11] determined the mass of droplets released when coughing. Depending on their method, they found 22.9 mg to 85 mg. The particles were distributed over a range from 5 μm to 300 μm (maximums at about 35 μm to 150 μm). In another study by Chao et al. [10], the particle size reached maximums of 6 µm (main peak) and 175 µm (secondary peak).

In sneezing fits, Han et al. [15] found two patterns of size distributions: unimodal in 12 subjects and bimodal in 10 subjects. For the unimodal distribution, the maximum was 341.5 µm to 398.1 µm. The geometric mean is 360.1 μm and the geometric standard deviation is 1.5 μm. For the bimodal distribution, the main peaks were at 72 μm and 386.2 μm with geometric standard deviations of 1.8 μm and 1.5 μm, respectively. Although only a very limited number of studies are available, the data shows that speaking produces smaller particles than coughing and sneezing.

The following data were measured for the initial air velocity during coughing: 11.7 m/s [10], 9.0 m/s [16] and 15.3 m/s for men and 10.6 m/s for women [17]. The air velocity during speaking was much lower with 3.9 m/s [10] and 4.07 m/s for men and 2.31 m/s for women [17]; respectively 1.08 m/s - 1.56 m/s for men and 1.53 m/s -1.64 m/s for women [18]. Tang et al. [19] determined maximum airflow velocities of 4.5 m/s for sneezing, 1.4 m/s for nasal breathing and 0.8 m/s for mouth breathing. The sneezing velocity obtained from this study was lower than in other studies. This could be due to different techniques used in different studies and the limitation of the imaging technique (shadowgraph imaging), which requires a temperature difference between the exhaled puff and the ambient air. Air velocity decreases exponentially with distance from the mouth [20].

### Use of face masks

The use of personalized protective equipment (e.g. face mask) is a common measure against the transmission of viruses worldwide. A distinction is made between three types: a) particle-filtering half masks (e.g. FFP2), b) medical masks of type I or II (e.g. surgical masks) and c) mouth and nose covers (e.g. fabric masks).

Several recent studies have measured the efficiency of face masks for trapping particles. Konda et al. [21, 22] tested the effectiveness of common cloth masks in the laboratory and found filtration efficiencies of 5% - 80% for particles <300 nm and 5% - 95% for particles >300 nm. However, their results are critically discussed, mainly due to the small pressure differences in this study compared to the pressure differences that occur under normal breathing conditions. This implies that the high filtration efficiency is mainly due to a very low air velocity of the particles [23, 24].

Other studies found lower efficiencies for surgical and cloth face masks at a realistic pressure drop and air velocities [25-28]. For example, Zhao et al. [28] found a filtration efficiency of 5% to 25% for common fabrics made of cotton, polyester, nylon and silk, a filtration efficiency of 6% to 10% for polypropylene spunbond and 10% to 20% for products paper base. Hill et al. [25] report a filtration efficiency of 17.4% for single layer cotton. On the other hand, the N95/PN95 masks evaluated by Hill et al. have shown significantly higher filtration efficiency (>98%) [25].

Surgical face masks can also reduce the transmission of coronaviruses and influenza viruses. Leung et al. [29] detected coronaviruses (human seasonal coronavirus, not SARS-CoV-2) in respiratory droplets and in aerosol samples without wearing face masks, but no viruses when wearing surgical face masks. In a model study, in which the filtration efficiency of a normal surgical mask for virus-laden aerosols was assumed to be 50%, an infection probability of <1% could be achieved even in a confined space.

Asbach et al. [8] have determined the filtration efficiency of different mask types as a function of particle size by means of electrical mobility analysis and also conclude that FFP2 masks are most effective when used properly.

### Risk of infection and modeling tools

The risk of viral infection in an enclosed space depends on various factors. These include the concentration and infection rate of the virus, the distance between people, personal protective measures and ventilation conditions. An effective process for reducing the concentration of particles in a room is dilution with clean and virus-free air [30, 31]. Outdoors, dilution is constantly taking place through natural air movement. Indoors, dilution can be achieved through efficient ventilation. If possible, windows should be opened and air movement should be ensured. The most effective way to do this is cross ventilation. The ventilation time required depends on the size of the room, the number and size of the windows and the temperature difference inside and outside. If necessary, the air exchange can be forced mechanically. Mobile air purifiers should only be used in exceptional cases.

The general risk of infection with SARS-CoV-2 viruses indoors has been addressed in various works. Buonanno et al. [32] compare different scenarios and activities (hospital, sports hall, public building, conference room or auditorium). Cortellessa et al. [33] examined the risk of infection as a function of the interpersonal distance, taking into account both the spatial and the temporal aspect. Qian et al. [34] analyzed 318 outbreaks in terms of location and social contacts. Deng and Chen [35] examined the risk of infection depending on the distance between people with and without masks. The authors conclude that if face masks are worn correctly, the distance between two people can be reduced to 0.5 m without increasing the risk of infection.

To calculate the risk of infection, many models require the so-called “quanta concentration” in the room air [36-38]. A quanta describes the amount of viruses that, if inhaled by a person, will lead to an infection with a given probability. The assumption is usually made that the infected and infectious persons are in the room at the same time and that the quanta concentration in the ideally mixed room air corresponds to the equilibrium concentration for the entire period. The quanta concentration can be calculated from the quanta emission rate. In general, quanta values can only be roughly estimated.

The risk of infection in a well-mixed room contaminated with virus-carrying aerosols is often estimated using the Wells-Riley equation (1). It is assumed that the risk of infection increases exponentially with the number of viruses inhaled. The Wells-Riley equation thus describes a classic one-zone model without considering concentration gradients.

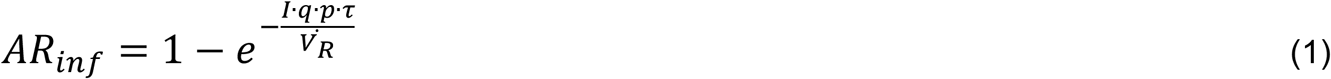

AR_inf_ is the absolute risk of infection. The probability that a person in the room will become infected under the given conditions, I is the number of infected people in the room, q is the quanta emission rate, p is the breathing volume flow of a person, Τ is the time that an uninfected person stays in the virus-polluted environment and 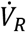 is the supplied air volume flow into the room.

The Wells-Riley approach was modified several times in order to be able to estimate the risk of infection under different conditions and for different scenarios [39-45]. The Wells-Riley model has the disadvantage that rapid and ideal mixing is assumed. Therefore, the risk of infection is the same at any point in the room. Guo et al. [46] have expanded the model for realistic room air flows, so that different infection risks can also be expected in different segments of the room. However, this is associated with a corresponding amount of computational effort.

Lelieveld et al. [47] developed a complex spreadsheet model containing a number of modifiable environmental factors with relevant physiological parameters and environmental conditions. The model takes into account the difference between everyday masks and FFP2 masks, different scenarios for air exchange, different virus mutations, so-called superspreaders and the change between quasi-stationary and transient conditions. The properties of the aerosols and viruses can be modified in detail. A similar model was published on the website of the International Laboratory for Air Quality and Health (ILAQH) at Queensland University of Technology (QUT) in Brisbane, Australia. However, both models have the disadvantage, just like the Wells-Riley approach, that they assume ideal mixing of the air. As already mentioned several times, this is usually only the case with simple supply air/exhaust air systems in small rooms. A multi-zone model for simulating the spread of infectious aerosols is available with the CONTAM software package [48].

The guideline concept of Bazant et al. [49] is based on models of airborne disease transmission. From this, the authors derive an upper limit for the cumulative exposure time. It is shown how this limit depends on the ventilation and air filtration rate, the dimensions of the room, the breathing rate, the respiratory activity and use of the face mask of its occupants, and the infectivity of the aerosols.

### Risk of infection at cultural events

Only a few works have been published on this topic to date. This may be due to the fact that the possible scenarios are varied and of a complex nature. In addition, rooms in which cultural events take place can only rarely be described with simple models. As a rule, complex approaches are necessary that take into account different concentrations in different areas (zones) of a building [48]. A typical dynamic scenario is shown in Figure 3.

**Figure 3.**
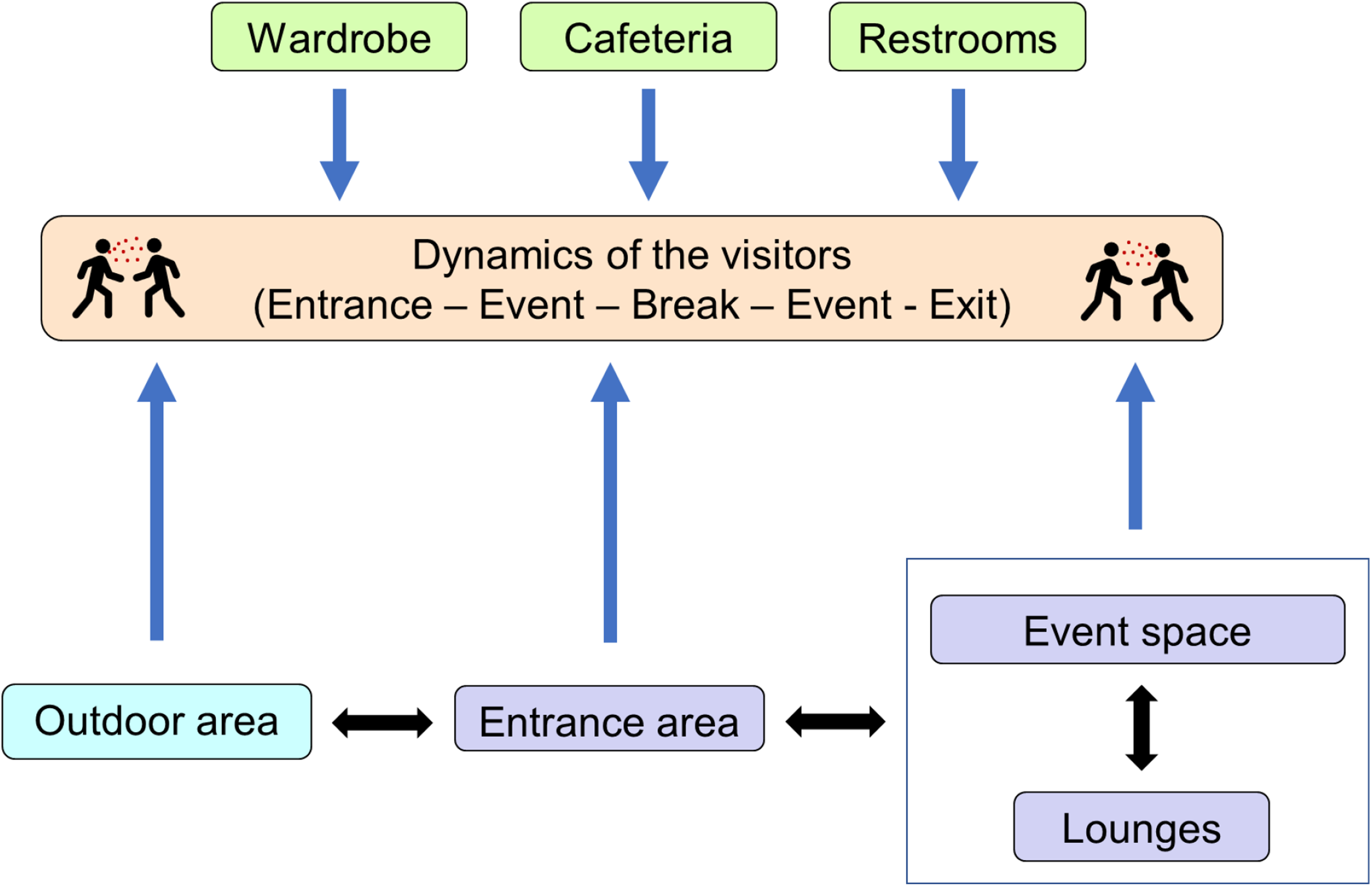
Dynamics of visitors during a cultural event.

The first phase of a cultural event is the arrival of the audience from admission to the start of the event. If there are assigned seats, the audience can be taken directly there, but it is also usual to stay in an entrance area first. There is often catering or the possibility of handing in outer clothing. The second phase is the event itself. In the simplest case, it is a cuboid room with a stage at the end. All listeners are on one level, the artists sit, stand or move in a slightly elevated position. Theaters usually have several levels (tiers) for the audience. The people are therefore at or above the level of the stage. Very large concert halls have grandstands, here the stage can also be in the middle. The third phase begins after the end of the event and ends when all guests have left the venue. Here it is common for people to remain in the event hall or in the entrance area for a while. Phases 1 and 3 can be controlled by appropriate logistics. However, there are often breaks during which visitors may move around the event complex. The published studies are discussed below. Table 1 provides an overview.

**Table 1.** Overview of published studies relating to the risk of SARS-CoV-2 infection at cultural events.

| Event | Type of study | Ref. |
| --- | --- | --- |
| Choir and other scenarios | Modeling study | [47] |
| Socio-cultural events | non-randomized controlled study | [61] |
| Chorus rehearsal | Estimation of infectious quanta | [50] |
| Wind instruments | Risk of spreading infectious particles | [51] |
| Concert hall | Evaluation of the infection risk using mannequins | [52, 53] |
| Indoor sports and cultural events | simulation study to estimate the burden of disease under conditions of controlled epidemics | [54] |
| Mass gatherings | Effectiveness of public health measures | [62] |
| Mass sporting and cultural events | Use of interviews and questionnaires, to describe the potential public health impact | [63] |
| Singing event | Evaluation of safer singing practices | [56] |
| Live concert | Randomized controlled trial of attendees | [57] |
| Indoor live concert | Screening of attendees for SARS-CoV-2 antigen before the event | [59, 60] |
| Concert events | Assessment of SARS-CoV-2 transmission using contract-tracing data | [55] |
| Carnival | Seroepidemiological observational study | [65] |
| Visitor's point of view | Online study with persons, who regularly attended events before the pandemic | [66] |
| Sport events | Assessment of the suitability and feasibility of a hygiene concept | [64] |

Lelieveld et al. [47] modeled the infection risk for four different indoor situations (office, classroom, choir, reception) and different scenarios (active and passive ventilation, type of masks, filters). It was assumed on the one hand that the exhaled number of viruses corresponds to the usual magnitudes (standard event) and on the other hand that the virus output is increased (superspreading event). In summary, choral singing caused the highest risks.

Miller et al [50] examined the spread of SARS-CoV-2 viruses at a church event. A total of 61 people met to sing in a choir, 53 of whom were infected. Precautions were taken during the rehearsal, including the use of hand sanitizer, no hugs, and no handshakes. All 120 chairs were arranged by three people who arrived early, and members sat in their usual chairs. Lateral spacing between chair centers (and hence nose-to-mouth spacing) was ~0.75 m, while front spacing between rows was ~1.4 m.

14 wind instruments were analyzed by Spahn et al. [51]. This was done qualitatively by making air currents visible while playing and then quantitatively by measuring the air speed at three distances (1 m, 1.5 m, 2 m) from the respective instrument. The measurements took place on musicians of the Bamberg Symphony Orchestra in their concert hall. The results show that while playing, no airflow was measurable beyond a distance of 1.5 m for any wind instrument, for brass instruments from the bell or for woodwind from the mouthpiece, vents or bell, regardless of volume, pitch or what was played. The air speed when playing corresponded to the value of 1 m/s that is usual in hall-like rooms. In the case of woodwind instruments, alto flute and piccolo, clear air movements could be observed near the mouthpiece.

Schade et al. [52] experimentally investigated the spread of aerosols in the audience area of several concert halls in order to assess their airway and thus the risk of spreading an infection with SARS-CoV-2. For this purpose, a dummy was used that emits simulated human breath with aerosols (mean diameter 0.3 mm) and CO_2_ with a horizontal exhalation speed of 2.4 m/s, measured 10 cm in front of the mouth. Aerosol and CO_2_ concentration profiles were determined using sensors placed around the dummy. No significant accumulation of aerosols and CO_2_ was detected at adjacent seats, provided that displacement ventilation was provided under each seat, which enabled a local fresh air vertical flow of at least 0.05 m/s, the air exchange rate was higher than 3 h^−1^ and the dummy wearing a surgical face mask. The experiments of Schade et al are described in more detail in another publication [53]. The different results depending on wearing or not wearing a face mask are also demonstrated here. Wearing a mask significantly reduces the risk of infection for people sitting in front of an emitter.

Moritz et al. [54] investigated the risk of droplet and aerosol transmission of SARS-CoV-2 during a mass event under three different hygiene practices and used the data in a simulation study to estimate the resulting disease burden under controlled epidemic conditions. The results show that the average number of measured direct contacts per visitor was nine people and that this can be significantly reduced through appropriate hygiene practices. A comparison of two ventilation variants with different air exchange rates and different air flows showed that the system with the worst performance caused a tenfold increase in the number of people exposed to infectious aerosols. The overall burden of infection from indoor mass accumulations therefore depends significantly on the quality of the ventilation system and hygiene practices. Assuming an effective ventilation system, the authors believe that indoor mass gatherings with proper hygiene practices have very little, if any, impact on the spread of the epidemic.

SARS-CoV-2 infection chains were investigated by Koizumi et al. [55] at several small concert events in the Osaka area, Japan.

Naunheim et al. [56] discuss the risk of infection with SARS-CoV-2 at singing events. The authors conclude that the risk can certainly be reduced by taking suitable measures, but that risks cannot be completely eliminated. Every community of singers and artists must do everything possible to mitigate the risk as much as possible, and then decide whether this risk reduction of SARS-CoV-2 transmission is sufficient to resume the singing activity under consideration of all the discussed factors. From today’s perspective, the statement sounds trivial, but leads to the conclusion that there can be no generally applicable recommendation. The measures must always be adapted to the respective situation.

Delaugerre et al. [57] compared the SARS-CoV-2 infection rates among those attending a large live concert on May 29, 2021 in Paris with those who were not present at that concert. They also rated the effectiveness of mask wearing. The results did not show a significantly increased risk of SARS-CoV-2 transmission among participants compared to non-participants. In this context, Schlagenhauf and Deuel [58] note that in May 2021 most SARS-CoV-2 infections in France were caused by the alpha variant. With the significantly more contagious delta variant (note: the delta variant refers to the time of writing of the article), the transfer of the results of Delaugerre et al. [57] to other conditions is not necessarily given.

Llibre and colleagues [59, 60] studied transmission rates during an indoor live music concert with 5000 people in Barcelona using antigen testing. Singing and dancing were allowed and no minimum distance was required. Six people who attended, none of whom were vaccinated, were diagnosed with COVID-19 within two weeks of the concert.

Ramos et al. [61] state that attendance to events that involved social interaction with a certified digital pass was not associated with an increased rate of SARS-CoV-2 infection compared to a control group. Logistical problems were infrequent and easy to solve, the participants’ overall opinion on the accreditation process was satisfactory.

Walsh et al. [62] have dealt with the risk of infection at mass events. Their review, taking into account 11 studies, found that implementing measures can reduce the risk of SARS-CoV-2 transmission. However, it is unlikely that this risk can be completely eliminated. All studies followed a multi-pronged, multi-measure approach. This seemed more effective than relying on a single measure. The number and intensity of measures implemented varied by study, with many being resource intensive. There is currently limited evidence for the effectiveness of measures to prevent SARS-CoV-2 transmission at mass gatherings. With such events resuming, continued known mitigation measures are required to limit the risk of transmission, as well as ongoing research and monitoring to assess the potential impact of these events on the wider population and healthcare system.

Smith et al. [63] used contact tracing data collected through telephone interviews and online questionnaires to investigate the different numbers of infections at sporting events with comparable numbers of visitors. These were attributed to socio-economic factors, different safety awareness among the visitors and their different dynamics of movement.

Geisler et al. [64] assessed the suitability and feasibility of a hygiene concept during a series of indoor sporting events. The increasing spectator numbers consolidated and optimized hygiene concept-related processes and demonstrated a high level of spectator compliance with the hygiene measures.

Streeck et al. [65] report a SARS-CoV-2 super-spreading event during carnival in a small German town. Due to quickly imposed lockdown measures and the resulting relatively closed community, this city has been considered a good model for studying infection dynamics. A seven-day observational study was conducted to collect information and biological samples from a randomized, household-based study population.

Another important point concerns the question of how visitors prepare for participation in an event, what preventive measures they take and what expectations they have of the organizer. This aspect was examined by Weber et al. [66] as part of an online study. The use of disinfectants was mentioned as the most important element of containment, followed by transparent information on the hygiene strategy, reduced crowds, optimized ventilation, body temperature measurement at the entrance, negative SARS-CoV-2 test, filling out a health questionnaire. Forgoing breaks and meals was seen as less important.

Vernez et al. [67] analyzed the situation in a poorly ventilated courtroom. There are certainly parallels to a cultural event here. The negotiation takes place on a kind of stage, with different people interacting with each other verbally. If necessary, the seats for spectators are separated from this stage. It was found that in such situations and without further protective measures, the risk of infection is particularly high.

### Criteria for ventilation conditions

The big problem with cultural and sporting events is the complexity of the halls and arenas. Only in a few cases will simple ventilation, as shown in Figure 4A, be sufficient. It must then be assumed that the air is completely mixed so that, as in school classrooms, the risk of infection can be assessed according to Wells-Riley.

**Figure 4.**
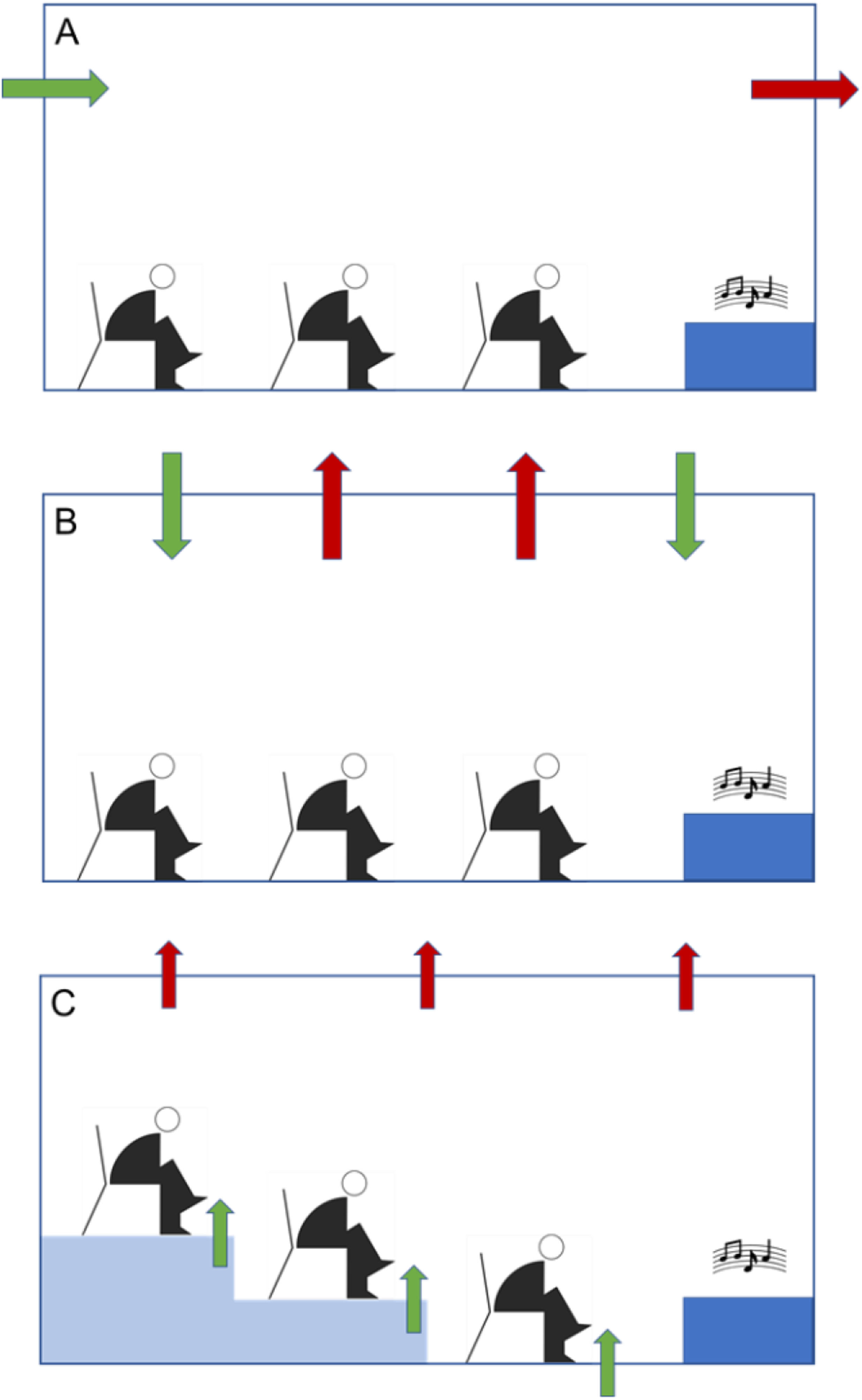
Different types of ventilation during a cultural event. A: simple ventilation by fans; B: ventilation system with inlet and outlet in the ceiling area; C: individual ventilation directly at the seat and outlet in the ceiling area. The green and red arrows indicate air inlet and outlet, respectively.

Larger halls often have multiple air inlets and outlets at the ceiling as shown in Figure 4B. Individual air supply makes sense for complex seating arrangements with several levels (see Figure 4C). However, this is only possible with full seating.

The example shown in Figure 5 is certainly at the limit of what is possible with the Wells-Riley equation. A room volume of V = 1260 m^3^ (21 m × 14 m × 5 m) was selected with the presence of N = 96 attendees and one infected person (I = 1) in the room. A respiratory volume flow of p = 0.54 m^3^/h and a residence time of *τ* = 2.5 h were assumed per person. The air exchange was between 0.1 h^−1^ and 10 h^−1^, the viral (infectious) release rate was q = 10 (green), 25 (red), 50 (blue) and 100 (yellow) quanta/h per person.

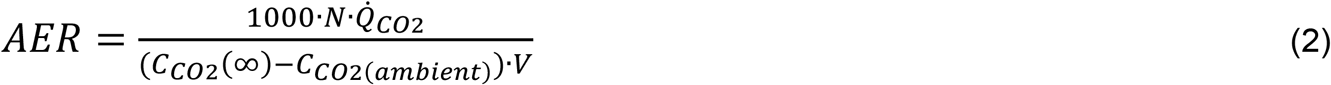

**Figure 5.**
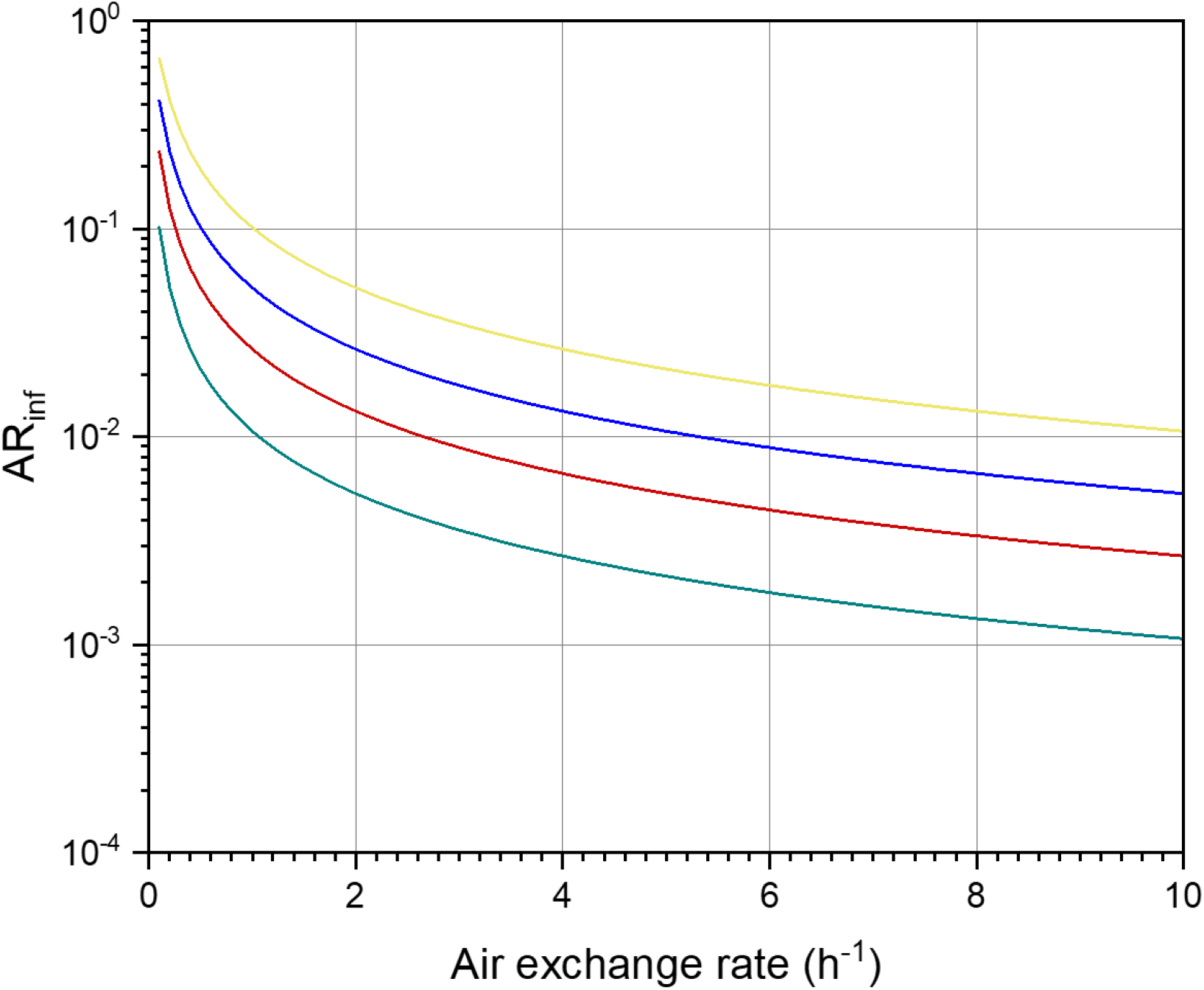
Absolute risk of infection AR_inf_ according to Wells-Riley depending on the air exchange rate. The calculation was carried out with equation (1) for different quanta, see text for parameters and colors of the curves.

It is further assumed that each person exhales 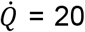 l of CO_2_ per hour and that the background concentration of CO_2_(ambient) is 400 ppm. According to Equation (2), the air change required for a CO_2_ steady-state concentration of <2000 ppm is 1 h^−1^. In order to permanently fall below a CO_2_ concentration of 1000 ppm, as a recommended indoor air concentration for example at schools [68] and rooms with many people inside [69], an air change of 2.6 h^−1^ would be necessary. Assuming that the distance regulation is observed (each person has 1.5 m × 1.5 m space), the theoretical absolute risk of infection AR_inf_ with an air change of 2.6 h^−1^ and the highest q is still 0.04. In practice, these values are likely to be far too optimistic. Carbon dioxide has a higher molecular weight and a higher density than nitrogen and oxygen and unless the air is well mixed it will tend to accumulate near the ground. This requires very good mixing conditions, which cannot be achieved with natural ventilation, and at the same time an increased air exchange to prevent contaminated air from being distributed in the hall.

Regardless of pandemics, it is generally important to ensure good ventilation conditions in rooms. This applies to cultural events as well as to school classrooms and means of transport. Corresponding measurements must be carried out with a high time resolution and should be automated. This usually excludes the direct detection of bioaerosols, so other parameters must be used. With regard to hygienic aspects, the carbon dioxide concentration in the room air serves as an indicator for air deterioration caused by human activity. The concept of the German Committee on Indoor Air Guide Values (AIR – formerly Ad hoc AG) [69] considers CO_2_ concentrations of less than 1000 ppm (0.1% by volume) to be hygienically harmless and CO_2_ concentrations greater than 2000 ppm to be hygienically unacceptable.

In rooms with high occupancy, the carbon dioxide concentration can serve as an indication of good or bad ventilation. Carbon dioxide has long been considered a reliable indicator of air exchange, a maximum CO_2_ concentration of 1000 ppm indicates hygienically sufficient air exchange under normal conditions. This means that information can be provided quickly and easily as to whether and when ventilation is necessary. However, this does not mean that a CO2 concentration of less than 1000 ppm generally protects against infection with SARS-CoV-2. Conversely, CO_2_ concentrations significantly or permanently greater than 1000 ppm indicate inadequate ventilation management with a potentially increased risk of infection. Inexpensive and reliable sensors are available for continuous carbon dioxide measurement. Their suitability could be demonstrated, for example, by measurements in local public transport [70, 71]. In the work of Querol et al. [71], the criterion is 800 ppm carbon dioxide (which is applied for example in European standards for the regulation of indoor ventilation systems in non-residential buildings) instead of 1000 ppm. Bauer et al. [72] also assess the air quality at singing events on the basis of 800 ppm CO_2_. Kitamura et al. [72] come to the conclusion that the regulation of supply and exhaust air based on CO_2_ measurements is an effective preventive measure against airborne infections at cultural events.

The determination of the particle concentration, especially PM_10_ and PM_2.5_, is also an important aspect for evaluating the indoor air quality [73, 74]. However, PM is not very suitable for checking ventilation requirements, since the dynamics of particles are already high due to the movement of people. Some of the particles are introduced via the outside air, some via clothing. Another parameter that can be measured easily and quickly is the sum of volatile organic compounds as the so-called TVOC_PID_ value [75]. TVOC stands for “Total Volatile Organic Compounds” [76] and PID for the measurement method using photoionization. However, TVOC_PID_ is not directly linked to the population density and the breathing rate of people, so that this parameter should at best be used in addition to the CO_2_ concentration.

### International recommendations and guidelines for cultural events

In June 2021, the European Commission published EU guidelines for the safe resumption of activities in the cultural and creative sectors [77]. Testing events organized in the EU have shown that few COVID-19 cases have been linked to transmission at or around cultural events. Particular requirements for participation and monitoring of data in the follow-up of these trials were essential for a safe organization. The Commission has developed population-related, individual-related and event-related indicators and recommends that the member states take these into account when opening up cultural activities.

The initiative of the German Federal Government has already been reported in an earlier chapter [1].

The Austrian guidelines of the Medical University of Vienna are intended to define medical framework conditions for the specific practice of cultural institutions or medical measures that are aimed at ensuring that visitors to a cultural institution are not exposed to a higher risk than with other contact with people in public space [78]. The focus here is on the complex of questions of the game operation with audience/visitors. Questions about presenting artists (actors, musicians/orchestras) as well as technical and administrative staff are addressed but not elaborated on.

In France, the Haut Conseil de la Santé Publique has published various texts on how to deal with the SARS-CoV-2 virus at cultural events.

In March 2022, the Australia Council of Arts published the “Audience Outlook Monitor” with recommendations on how to behave when attending live events. Website: https://www.thepat-ternmakers.com.au/blog/2021/audience-outlook-monitor-march-2022-key-findings, accessed on 02.07.2022.

In the United States, the Centers for Disease Control and Prevention (CDC) issues guidelines for small and large gatherings and events, which are updated regularly. Website: https://www.cdc.gov/coronavirus/2019-ncov/your-health/gatherings.html, accessed on 07.02.2022.

## Conclusions

Cultural events take place under very different conditions. It is therefore practically impossible to predict the exact risk of infection for a specific situation or to derive general rules that go beyond the known measures of vaccination, testing, masks and distance. In addition, there are a number of important factors that cannot be controlled or regulated. In European countries, the vaccination rate varies greatly from region to region, and counterfeit vaccination and test certificates must also be expected. Furthermore, the emotions of the viewers must be taken into account. It is certainly difficult not to move or not to sing along in an appealing concert. For very small events with up to approx. 50 participants and seated people, the criteria for classrooms can be created if necessary. Such simple scenarios can be estimated using the Wells-Riley equation or similar models. Adequate mixing of the air can already be achieved here with simple ventilation systems.

Very large halls with up to 20,000 seats for visitors are just as difficult to model as architecturally complex medium-sized concert halls. However, the event itself is only one aspect. Equally important are questions of guiding people when entering and leaving and the connection to other premises (VIP lounges, cafeteria, etc.). Whatever measures are taken by the organizers, successful implementation always requires a certain understanding and awareness of the situation. All of these points make it difficult to collect meaningful data at a cultural event, with uncertainties increasing as the number of participants increases. In addition, the infectious persons are not necessarily recorded representatively in test series on site. It is less likely that people who are generally critical of vaccinations and tests will take part in a study on site.

In general, it is not possible to base a risk analysis on a single parameter. For example, the distance control or staggered seating arrangements will be of little use if the ventilation in the hall is insufficient. Table 2 gives a selection of the main influencing variables. The most important point is definitely air exchange. Sufficient uncontaminated fresh air must be supplied to the room, which is appropriate for the room size and the number of people in the room. This fresh air must be effectively distributed in the room. This requires effective concepts for both the introduction and the air flow, which are adapted to the individual room or hall.

**Table 2.** Parameters that should be considered when holding a cultural event under pandemic conditions.

| Parameter | Implementation/performance |
| --- | --- |
| Room type | Floor space and room height<br>Spatial geometry of the room<br>Number of levels<br>Arrangement of the levels (if several exist)<br>VIP lounges available?<br>Position of stage (if any) |
| Dynamics before, during and after the event | Distance rules at the entrance and exit<br>Duration of the event<br>Breaks<br>Catering |
| Type of performance | Reading, lecture<br>Play (language only)<br>Operetta, opera<br>Concert (only instrumental or with vocals) |
| Position of the audience | Standing<br>Sitting (side by side, staggered, others) |
| Reactions of the audience | Sitting or standing still<br>Moving or dancing<br>Singing along |
| Vaccination status and behavior of the audience | Vaccinated (1 x, 2 x, 3 x, more...)<br>Negative test<br>Masks (FFP2, other medical masks)<br>Use of disinfectants |
| Room ventilation | Natural ventilation (opening windows and doors)<br>Fans (without heat exchanger)<br>Ventilation system (number and position of air inlets and outlets)<br>Air volume flow and proportion of recirculated air<br>Personalized supply air<br>Connection of the ventilation system to other rooms, e.g. B. VIP lounges |
| Air cleaning measures | None<br>Integrated into the ventilation system<br>Mobile air purifiers<br>Spraying of active substances |

Simple systems, such as an air inlet and an air outlet, will only make sense in relatively small classroom-sized rooms. More complex systems are required for events with many participants. In large arenas, it makes sense to individually adjust the air supply to the room or room use (event hall, lounges, cafeteria, entrance area, etc.). In addition, measuring the carbon dioxide concentration at several points in the room is a very helpful parameter for assessing the need for ventilation and airflow. It has already been mentioned that there is hardly any experience and data on the risk of infection with SARS-CoV-2 viruses at cultural events, which makes it even more difficult to develop the necessary measures.

The use of mobile air cleaners and similar disinfecting measures at cultural events will only be possible and meaningful in exceptional cases. Uhde et al. [79] provide an overview on this topic with reference to classrooms. It is important that a mobile air purifier cannot replace the necessary fresh air supply.

Finally, it must be clearly pointed out once again that any recommended action for hygiene and ventilation measures in cultural institutions under pandemic conditions can minimize the risk of infection, but cannot completely rule it out. The measures planned by the organizers can only relate to possible exposure to infectious aerosols via the airway. It has to be assumed that the visitors protect themselves individually and also keep the recommended or specified distances. The latter point in particular is likely to be essential, as analyzes of passenger flows in local and long-distance public transport have shown.

## Data Availability

All data produced in the present study are available upon reasonable request to the authors

## Acknowledgements

Tunga Salthammer is grateful for the financial support from the Staatsministerin für Kultur und Medien (BKM) der Bundesregierung, Berlin, Germany, WKI project no. 11-02657-2240-00300.

## Author’s contributions

TS and HJM equally contributed to the design of the study and the evaluation of the results. TS took the lead in writing and designed the figures. Both authors read and approved the final manuscript.

